# The genetic architecture of Alzheimer disease risk in the Ohio and Indiana Amish

**DOI:** 10.1101/2021.07.06.21259932

**Authors:** Michael D. Osterman, Yeunjoo E. Song, Larry D. Adams, Renee A. Laux, Laura J. Caywood, Michael B. Prough, Jason E. Clouse, Sharlene D. Herington, Susan H. Slifer, Audrey Lynn, M. Denise Fuzzell, Sarada L. Fuzzell, Sherri D. Hochstetler, Kristy Miskimen, Leighanne R. Main, Daniel A. Dorfsman, Paula Ogrocki, Alan J. Lerner, Jairo Ramos, Jeffery M. Vance, Michael L. Cuccaro, William K. Scott, Margaret A. Pericak-Vance, Jonathan L. Haines

## Abstract

Alzheimer disease (AD) is the most common type of dementia and is currently estimated to affect 6.2 million Americans. It ranks as the sixth leading cause of death in the United States and the proportion of deaths due to AD has been increasing since the year 2000 while the proportion of many other leading causes of deaths have decreased or remained constant. The risk for AD is multifactorial, including genetic and environmental risk factors. Though *APOE* remains the largest genetic risk factor for AD, more than 26 other loci have been associated with AD risk. Here, we recruited from a population of Amish adults from Ohio and Indiana to investigate AD risk and protective genetic effects. With slightly lower incidence and later age of onset, it is thought that the Amish may hold protective genetic variants for AD. As a founder population that typically practices endogamy, variants that are rare in the general population may be at higher frequency in the Amish population. We characterized the genetic architecture of AD risk in the Amish and compared this to a non-Amish population, elucidating the lower relative importance of *APOE* and differing genetic architecture of the Amish compared to a general European ancestry population.

## INTRODUCTION

Alzheimer disease (AD), the most common type of dementia, is the sixth leading cause of death in the United States and occurs in over 35% of individuals age 85 and older.^1,2^ It is currently estimated that 6.2 million Americans are living with AD.^1^ Deaths attributable to AD increased by 146.2% from the years 2000 to 2018 whereas other leading causes of death remained constant or decreased.^1^ This burden of AD is expected to increase in the coming years due to increased longevity and decreased fertility, known as population aging.^1,3,4^ The cost of managing AD will continue to increase with an expected annual global cost surpassing $50 billion by 2050.^1,5,6^ People living with AD also suffer from severe degradation of their quality of life including reduced independence and being at higher risk of somatic and psychiatric comorbidities.^7–9^ Improved understanding of AD risk and subsequent improvements to screening, prediction, and prevention efforts are needed to reduce these burdens. As current medications only marginally and temporarily delay the progression and lessen the severity of AD, its growing prevalence serves as an imperative issue.

Risk for AD is multifactorial, including genetic and environmental risk factors.^8,10,11^ While only 2-5% of all cases of AD are strongly familial (e.g. result from high penetrance mutations),^12^ the overall heritability of late-onset AD is estimated to be as high as 70% based on twin studies and genome-wide association studies (GWASs); however, such estimates can vary by population and environment.^13–15^ Genetic risk for AD is complex, including more than 26 independent associated loci spanning diverse population groups.^16–18^ Despite this large number of loci, the currently confirmed loci associated with AD risk account for only a small proportion of the overall heritability of AD.^15,16^ Increased sample sizes and diversity of study populations will help GWASs to elucidate the remainder of the heritability.

The largest genetic risk for AD is conferred by the apolipoprotein E (*APOE*) locus^19^ on chromosome 19 with 3 to 15 times increased risk for those holding either one or two copies of the *e4* risk allele compared to those holding no risk alleles.^20^ This association between AD and *APOE* has been replicated across many different and diverse populations.^21–23^

One such population is the Amish: descendants of German and Swiss Anabaptist immigrants who settled in the United States during the eighteenth and nineteenth centuries. Communities currently living in Holmes County, Ohio and Elkhart & LaGrange Counties, Indiana are mostly descendants from the German Palatinate, while the communities in Adams County, Indiana largely descend from Swiss Anabaptist immigrants.^24–26^ The expansion of the Amish from a population of less than 1,000 founders in the United States^27^ with subsequent cultural and religious isolation has restricted the introduction of new genetic variation. This leads the Amish to be representative of a subset of a more general European gene pool. Because of these factors, the Amish are a unique population that can serve as an ideal candidate for genetic research. Due to endogamy, some variants rare in the general European population may be at higher frequency in the Amish, allowing for detection and consideration of effects that may not otherwise be captured in studies of the general population.^28^ This situation is ideal for investigation of susceptibility genes for complex traits, including AD.

A slightly lower prevalence of AD has been reported within Amish populations, even after accounting for the effect of a lower frequency of the *APOE e4* risk allele.^29–31^ Improved understanding of what protective or other risk-bearing variants the Amish may be enriched for could prove helpful in improving general understanding of genetic risk of AD.

We have recruited adults from Amish families living in Holmes County, Ohio and Elkhart, LaGrange, and Adams Counties, Indiana. Our current focus is to recruit individuals who are cognitively unimpaired relative to age-normed benchmarks (CN) but at elevated risk for developing AD. We characterize this population and compare with a non-Amish European-ancestry population living in the US for age, *APOE* genotype, and both a genetic risk score (GRS) using genome-wide significant variants and a polygenic risk score (PRS) spanning the entire genome.

## METHODS

### Subjects

Individuals included in this study have been recruited over the past 20 years for multiple studies of AD or dementia,^29,32–34^ age-related macular degeneration,^35–37^ and successful aging.^38– 40^ For all studies, the primary criteria for enrollment included being age 50 or older, being part of the Amish community, and being of Amish descent. All individuals were screened for cognitive status. In addition, for the current study, individuals were enrolled if they were known not to be cognitively impaired (CI) and were age 76 and older. We prioritized enrollment of individuals with at least one family member with probable or confirmed AD. Participants were recruited from Amish families living in Holmes County, Ohio and Elkhart, LaGrange, and Adams Counties, Indiana.

### Cognitive Screening

Depending on the specific study, at time of enrollment, individuals were cognitively screened using a combination of the 3MS education-adjusted examination (all individuals),^41^ the AD8 checklist,^42^ the CERAD word list learning test,^43^ and the Trail Making test (for the Alzheimer disease and successful aging studies).^44^ Individuals were classified as CN or CI based on established cutoffs.^41,44^ Individuals initially classified as CI were further evaluated by a clinical adjudication board, comprised of neurologists and neuropsychologists, to further classify them as having mild cognitive impairment (MCI), AD, cognitive impairment, not dementia (CIND), having an unclear status.

### Genotyping

At time of enrollment, 30 milliliters of blood were collected from all participants for use in direct DNA extraction and storage of plasma. Genotype data were collected using an Illumina Expanded Multi-Ethnic Genotyping Array^45^ with custom content (MEGAex+3k) or an Illumina Global Screening Array^46^ (GSA). The MEGAex chip includes over 2 million markers whereas the GSA chip includes a base quantity of 660,000 markers. When performing chip genotyping, we also included customized content of up to 6,000 variants to the MEGAex chip, including over 1,100 novel varaints that have already been identified from our previous Amish whole exome sequencing (WES) and whole genome sequencing (WGS) studies and other associated variants from GWAS and the National Institute on Aging’s Alzheimer’s Disease Sequencing Project^47,48^ (ADSP) studies that are not already on the chip. After genotype data were attained, imputation was performed based on a Haplotype Reference Consortium (HRC) panel.^49,50^ We investigated genetic relationships of individuals within the overall study population by calculating kinship coefficients using KING 2.26.^51^ Further, we compared the average genetic relationship across subpopulations based on recruitment site and cognitive status.

### Quality Control

Quality control (QC) was performed on MEGAex+3k and GSA genotyping chip sets independently, with each containing samples from both Indiana and Ohio. A total of 774 individuals in the Illumina MEGAex+3k array met the QC threshold of 3% for genotype missingness. There were 1,973,806 SNPs in this initial set of autosomal and X chromosome SNPs. All SNPs genotyped in < 5% of the individuals (n=52,393) were dropped. Additionally, monomorphic (n=1,235,890) and duplicate (n=1,471) SNPs were excluded. Common SNPs (MAF >=1%) were evaluated for deviation from Hardy-Weinberg Equilibrium (HWE) and dropped if the p-value was < 1 × 10^−6^ (n=5,518). Mendelian error checking was performed on the related individuals within the set and any identified genotypic errors were zeroed out for all members of the affected family. Missingness and HWE were repeated after Mendelian error checking. The final, cleaned MEGAex+3k data set consisted of 774 individuals and 655,441 SNPs (chromosomes 1-22, X).

1,322 individuals had < 3% missing genotypes for the Illumina GSA array. Of the 703,560 genotyped SNPs, 1,470 were genotyped in < 10% of the individuals and were dropped. Additionally, monomorphic (n = 144,333) and duplicate (n = 4,656) SNPs were excluded. Common SNPs (MAF >= 1%) were evaluated for deviation from HWE and dropped for p-values < 1 × 10^−6^ (n = 847). Mendelian error checking was performed on the related individuals within the set and any identified genotypic errors were zeroed out for all members of the affected family. Checks for missingness and HWE were repeated after Mendelian error checking. The final, cleaned GSA data set consisted of 1,322 individuals and 545,470 SNPs (chromosomes 1-22, X).

Imputation was run on the Michigan Server using the HRC reference set. The MEGAex+3k and GSA data sets were imputed separately, and each was submitted using the GRCh38 build for autosomes and hg19 for the X chromosome. The reference population for HRC was European and the phasing was done using the Eagle option. Each data set underwent quality control separately after imputation. For rare SNPs (MAF < 0.01) an INFO score minimum of 0.8 was required. Common SNPs were considered to have passed QC with an INFO score of 0.4 and above. The MEGAex+3k set had 1,059,138 rare (MAF < 0.01) SNPs that passed QC and 7,722,065 common SNPs. The GSA dataset had a total of 1,423,947 rare SNPs that passed QC and 7,777,352 common SNPs that met the threshold. The two separately imputed sets were then merged after QC into one set of 2,096 samples using overlapping SNPs contained in both. The final imputed dataset contained 8,311,803 SNPs. Of these, 759,280 were rare and the remaining 7,552,523 were common.

We compared the Amish population to an existing source of non-Amish, European-ancestry individuals living within the US. The ascertainment for this population has been described elsewhere.^52,53^ This population included individuals ascertained from the University of Miami at the John P. Hussman Institute for Human Genomics, the Vanderbilt University Center for Human Genetics Research, and Duke University. After standard quality control, a total of 2,470 adults were included with an approximate 1:1 case-control ratio. Case status was determined by autopsy when possible. Otherwise, diagnoses were evaluated by two independent neurologists. Other phenotype information includes sex, age of exam, and age of onset in cases.

### Comparisons in Genetic Risk of AD

The Amish population and comparison group were initially compared for distributions of sex, age, and cognitive status. Comparisons by genetic risk factors were performed in subsets of the overall population after exclusion of individuals under age 75 years old to account for the late age of onset of AD^30^ in addition to differences in age distribution between the Amish data and the non-Amish comparison data (**Supplemental Figure 1**).

The GRS was generated using 31 genome-wide significant variants, excluding *APOE* variants, as reported in the recent Jansen et al. (2019)^17^ genetic meta-analysis. The GRS was constructed using PRSice-2^54^ and goodness of fit was assessed in R version 3.5.1.^55^ Dosage information was considered for imputed SNPs. For ease of interpretation, the mean and standard deviation of the GRS were scaled to zero and one, respectively.

The PRS was generated using a pruning and thresholding approach in PRSice-2^54^ and the best-fit PRS model, in terms of correlation coefficient *R*^*2*^, across the combined Amish and non-Amish dataset was used. All SNPs from the Jansen et al. (2019)^17^ meta-analysis were included for PRS construction except for those within 500 kilobases of either *APOE* SNP (rs429358 and rs7412). The parameters for clumping in the construction of PRSs included a 500 kilobase window centered on each index SNP and an *r*^2^ threshold of 0.1. Dosage information was considered for imputed SNPs. A best-fit PRS was chosen in combined data after applying across different potential p-value thresholds of included index SNPs. For ease of interpretation, the mean and standard deviation of PRS were scaled to zero and one, respectively.

Distributions of the GRS and PRS were compared across the populations and by Alzheimer disease or other dementia case status. GRS and PRS models were compared with an *APOE*-only model, covariate-only (sex and age) model, and a combined *APOE* and covariate model. Additional models were constructed including GRS and PRS to investigate overall predictive ability of the risk scores with and without the presence of the other variables. The predictive value of the constructed models was assessed by area under the receiver operating characteristic (ROC) curve (AUC).

## RESULTS

After quality control and assurance, the genotype information of 2,096 Amish individuals was available for analysis. Of these, 1,965 had a cognitive exam performed. The final population included 1,146 females and 819 males (**Table 1**). Of these, 1,367 were classified after consensus expert review as CN, 385 were CI, 18 had mild cognitive impairment (MCI), and 326 were unclear or missing (Table 1). Among the 385 with CI, 152 individuals (7.3% of the total sample) were considered to have probable or confirmed AD or other type of dementia. The mean and median age of the Amish population sample were 75.17 and 79, respectively, with a range of 21 to 110 years old. This includes 1,198 individuals of age 75 years old or older. After exclusion of individuals under age 75 years, *APOE* genotypes for the Amish and comparison groups demonstrate a lower prevalence of *e4* alleles and higher prevalence of *e2* alleles in affected Amish individuals than non-Amish cases (**Table 2**). The unaffected Amish have a similar distribution of *APOE* genotype to that of the non-Amish controls, except for a lower prevalence of the *e2*|*e3* genotype.

**Table 1.** Demographic information and cognitive status of Amish study population and non-Amish comparison group. *\*In the Non-Amish population, participants’ cognitive status was characterized as either AD or CN, whereas in the Amish population, participants were sub-divided into additional classifications of cognitive status, including CN, MCI, Borderline, or CI (of which “AD or other dementia” was a subset of all CI individuals). **Other category includes individuals that are cognitively impaired through other mechanisms, including Parkinson’s Disease and stroke. Abbreviations: CN = cognitively normal, MCI = mild cognitive impairment, AD = Alzheimer disease*.

| Trait | Amish n (%) | Non-Amish n (%) |
| --- | --- | --- |
| Female | 1,146 (58.3) | 1,449 (62.9) |
| Age 75+ | 1,198 (61.0) | 960 (52.5) |
| Cognitive Status* |  |  |
| CN | 1,367 (69.6) | 1,126 (48.9) |
| MCI | 18 (0.9) | - |
| CI | 385 (19.6) | 1,177 (51.1) |
| AD or other dementia | 152 (7.3) |  |
| Unclear | 189 (9.7) | - |
| Other** | 5 (0.3) | - |

**Table 2.** *APOE* distribution by population and AD case status of individuals with known *APOE* genotype. A p-value for two-sample population proportion Z-score test in individuals age 75 years and older is provided for comparisons between affected Amish vs. non-Amish cases in addition to unaffected Amish vs. non-Amish controls. An asterisk (*) denotes a significant difference in proportion at α = 0.05.

| *APOE* Genotype | Affected Amish n (%) | Non-Amish cases n (%) | p-value | Unaffected Amish n (%) | Non-Amish controls n (%) | p-value |
| --- | --- | --- | --- | --- | --- | --- |
| *e2\|e2* | 0 (0.0) | 0 (0.0) | - | 1 (0.1) | 3 (0.7) | 0.052 |
| *e2\|e3* | 14 (10.2) | 21 (3.9) | 0.003* | 84 (9.0) | 52 (12.7) | 0.038* |
| *e2\|e4* | 2 (1.5) | 13 (2.4) | 0.503 | 15 (1.6) | 12 (2.9) | 0.112 |
| $e3\vert e3$ | 70 (51.1) | 201 (37.1) | 0.003* | 632 (67.4) | 264 (64.2) | 0.258 |
| $e3\vert e4$ | 42 (30.7) | 240 (44.3) | 0.004* | 195 (20.8) | 78 (19.0) | 0.447 |
| $e4\vert e4$ | 9 (6.6) | 67 (12.4) | 0.055 | 11 (1.2) | 2 (0.5) | 0.234 |
| Total $e2$ | 16 (5.8) | 34 (3.1) | - | 101 (5.4) | 70 (8.5) | - |
| Total $e3$ | 196 (71.5) | 663 (61.2) | - | 1543 (82.2) | 658 (80.0) | - |
| Total $e4$ | 62 (22.6) | 387 (35.7) | - | 232 (12.4) | 94 (11.4) | - |

### Relatedness

Average kinship coefficient across all individuals in the Amish study population was calculated to be 0.003703 which is equivalent to between third and fourth cousins. Average kinship coefficient across subpopulations by primary study site and CI status were similar (**Supplemental Table 1**).

### Genetic Risk Score

After GRS construction, we observe, in general, less variance among Amish GRS, regardless of affection status, than in the non-Amish comparison group (**Figure 1**). Though the mean and median GRS are greater for the affected Amish than in the unaffected Amish, this difference is not statistically significant at α = 0.05. Further, the Amish population has no individuals among the 13 highest values of the GRS in the combined analysis, regardless of affection or case status.

**Figure 1.**
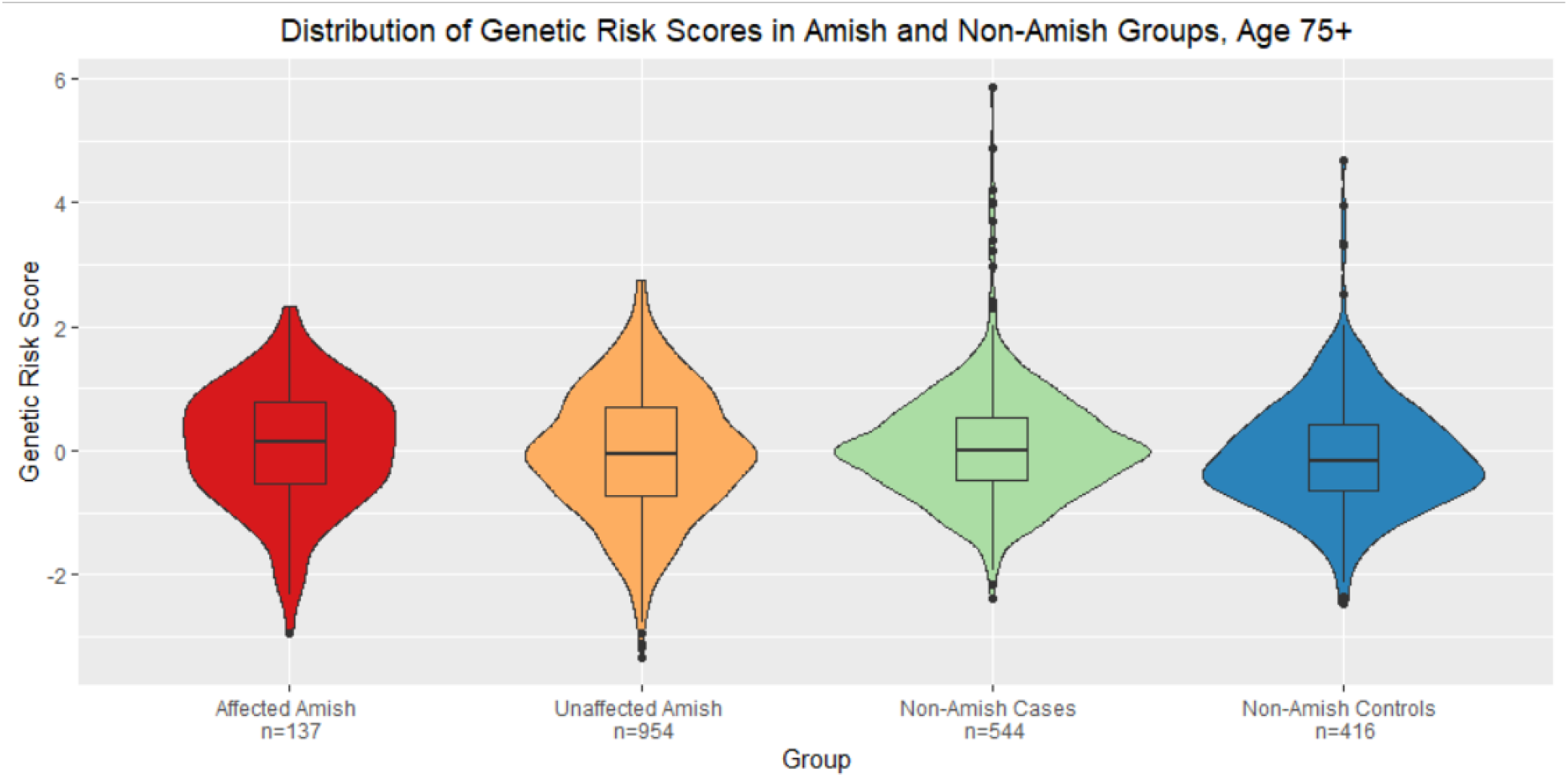
Distribution of Genetic Risk Scores by Amish and Alzheimer Disease Status. Genetic risk scores were constructed using only genome-wide significant single nucleotide polymorphisms. Only individuals age 75 years or older were included. GRS was able to distinguish (p=0.013) between the non-Amish cases and controls but not between the Amish affected and unaffected nor the Amish affected and non-Amish controls.

### Polygenic Risk Score

We observed that the values of PRS in the affected Amish individuals are lower than in the non-Amish cases. The values of PRS in the non-Amish controls are generally lower than that of the unaffected Amish. Overall, the difference in PRS values between the Amish affected and unaffected is much smaller than between the non-Amish cases and controls. The 29 highest PRS values all belonged to non-Amish individuals with the 13 highest of these belonging to non-Amish cases. PRS was unable to distinguish between affection status in the Amish (p = 0.7) but was able to distinguish between case status in the non-Amish population (p < 0.0001). PRS was also able to distinguish (p < 0.0001) between affected Amish and non-Amish controls in addition to non-Amish cases and unaffected Amish (p < 0.0001).

**Figure 2.**
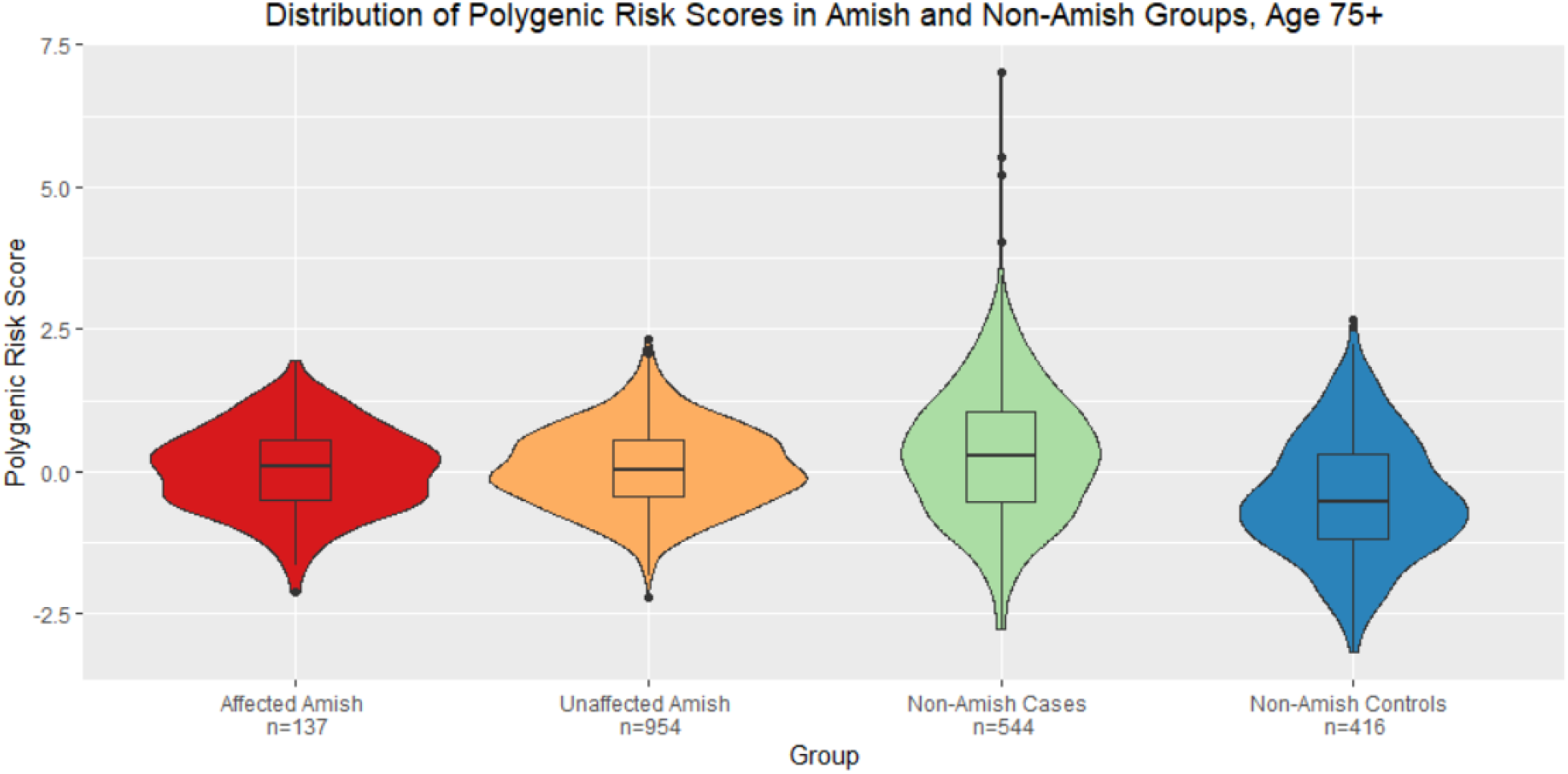
Distribution of Polygenic Risk Scores (PRS) by Amish and Alzheimer Disease Status. Polygenic risk scores were constructed using a pruning and thresholding approach on all variants, excluding those within 500 kilobases of either *APOE* single nucleotide polymorphism. Only individuals age 75 years or older were included.

We evaluated the association of the GRS, PRS, sex, age, and *APOE* genotype with the primary outcome of AD or other dementia by building a series of logistic regression models, after stratification by source population. Age was associated with the primary outcome across all models at α = 0.05.

We found that none of the *APOE* genotype categories are associated with affected vs. unaffected status in the Amish whereas each of the *APOE* genotype categories including at least one *e4* allele were associated with case status in the non-Amish population at α = 0.1 (*e2*|*e4* p-value = 0.062; *e3*|*e4* p-value = 0.004; *e4*|*e4* p-value = 0.0003).

The GRS and PRS were associated (p < 0.05) with the primary outcome across all tested models including PRS in the non-Amish populations. However, GRS and PRS were not significantly associated with the primary outcome in the Amish population, despite having an odds ratio (OR) > 1 across all models including GRS and PRS.

We also evaluated goodness of fit through AUC across each of these models (**Table 3**). We determined that the AUC of the sex and age only (covariate) model is larger in the Amish (0.693) than the non-Amish population (0.601). By contrast, we determined that the AUC for an *APOE* genotype only model is larger in the non-Amish population (0.712) than in the Amish population (0.594). The GRS models performed similarly in the Amish and non-Amish populations. A higher AUC was observed for the PRS models in the non-Amish population than in the Amish population.

**Table 3.** Goodness of fit of predictive models by sex, age, *APOE* genotype, genetic risk score, and polygenic risk score. For each constructed logistic regression model, area under the curve of a receiver operating characteristic curve is presented. The outcome of interest in each model is probable or confirmed Alzheimer disease or other dementia. *Abbreviations: COV = Sex and age covariates; GRS = genetic risk score including only genome-wide significant single nucleotide variants, excluding APOE variants; PRS = polygenic risk score using a pruning and thresholding approach, excluding single nucleotide polymorphisms within 500 kilobases of APOE variants*.

| Group | COV | *APOE* | COV + *APOE* | GRS | GRS + *APOE* | GRS + COV | GRS + COV + *APOE* | PRS | PRS + *APOE* | PRS + COV | PRS + COV + *APOE* |
| --- | --- | --- | --- | --- | --- | --- | --- | --- | --- | --- | --- |
| Amish | 0.693 | 0.594 | 0.741 | 0.534 | 0.608 | 0.695 | 0.744 | 0.506 | 0.595 | 0.693 | 0.741 |
| Non-Amish | 0.601 | 0.712 | 0.765 | 0.551 | 0.731 | 0.613 | 0.769 | 0.687 | 0.788 | 0.710 | 0.812 |

## DISCUSSION

This study characterized and evaluated the genetic risk for AD in an Amish population and compared it to a non-Amish population of predominantly European ancestry. The results indicate that there exists not only less variation in *APOE* genotype within the Amish, but also that *APOE* genotype may not play as large of a role in development of AD or other dementia as within a typical European ancestry population. Our results support the notion that *APOE* has a smaller effect on AD risk in the Amish population than in a non-Amish population,^30^ possibly due to the lower prevalence of *APOE e4* in the Amish population.

Non-*APOE* GRS and PRS have only moderate predictive value on their own but in addition to covariates, they do provide a meaningful increase in predictability in a logistic regression model for case/affected status. We determined that, based on a GRS of genome-wide significant SNPs from a recent meta-analysis of GWASs,^17^ there exists more variation among genetic risk in a non-Amish population than in an Amish population. When extending to a PRS analysis, this phenomenon is much more prominent. The PRS model also added additional distinguishing ability in AD or other dementia status in the non-Amish population. We also determined that a non-*APOE* GRS and PRS do not seem to differ greatly between affected Amish and non-Amish cases, suggesting that risk scores created using effect size weights derived from non-Amish European samples may not accurately predict risk in the Amish. This is somewhat similar to previous findings^29^ of GRSs that included *APOE* but highlights that *APOE* still plays an important role in AD prediction in the Amish.

In predicting the primary outcome of AD or other dementia, our results suggest that age is the most crucial risk factor in the Amish population whereas *APOE* and PRS bear greater importance in the non-Amish population. We observe much worse predictive ability when using a PRS that includes SNPs that do not meet genome-wide significance criteria in the non-Amish population compared to the Amish population, suggesting that the underlying genetic architecture for AD risk is dissimilar to that of a general European ancestry population, especially among SNPs that do not meet criteria for genome-wide significance in the non-Amish population.

The lower prediction ability in the Amish for GRS and PRS comprising known AD risk factors suggests that the risk profile in the Amish is significantly different – either through variation in the effect size for these known alleles, the existence of unidentified AD risk factors, or, likely, both. When combining this with information that the Amish have lower prevalence of cognitive impairment and dementia,^29,30,56^ it becomes clear that between their genetic risk factors and lifestyle, the Amish are somewhat protected from these outcomes in a way that using risk estimates from a general European ancestry population cannot explain. This warrants further investigation as the Amish are a sub-population of European immigrants that have practiced endogamy since arriving in the United States. Our results add to mounting evidence that there is genetic risk in the Amish that is not captured by genetic risk scores derived from non-Amish populations.

We conclude that there are evident differences in the genetic architecture for AD risk in the Amish compared to a non-Amish European ancestry population, especially in terms of *APOE* distribution, PRS distribution, and their conferred risk. Future genomic studies including the Amish should consider using effect estimates from an Amish analysis to determine if there are substantial differences in predictive ability than are seen after PRS construction using effect estimates from a non-Amish population. Identification of why the Amish appear to be relatively protected from AD and cognitive impairment, in general, warrants further study to identify risk factors enriched in the Amish that may enhance previously identified pathways important in the development of AD and identify additional pathways or mechanisms that contribute to or protect against cognitive decline. By extending this cohort through new recruitment and longitudinal follow-up, the power of this cohort to identify both novel risk and protective genetic loci, and potential predictors of progression from normal to AD will be increased. This will allow for better detection of rare effects and better understanding of the differences in the genetic risk of AD between the Amish and non-Amish populations.

## Supporting information

STROBE Checklist

## Data Availability

Data will be made available through NIAGADS (https://www.niagads.org/) upon publication.

## ACKNOWLEDGEMENTS

We thank the Amish families for their willing participation in our study. We used the Anabaptist Genealogy Database and Swiss Anabaptist Genealogy Association. This study is supported by National Institutes of Health / National Institute on Aging, grant AG058066 (to Jonathan L. Haines, Margaret A. Pericak-Vance, and William K. Scott). Finally, we acknowledge the resources provided by the Department of Population and Quantitative Health Sciences, School of Medicine at Case Western Reserve University and the John P. Hussman Institute for Human Genomics at University of Miami, Miller School of Medicine.

## REFERENCES

1. 2019 Alzheimer’s disease facts and figures. Alzheimer’s Dement. 15, 321–387 (2019).

2. Hebert, L. E., Weuve, J., Scherr, P. A. & Evans, D. A. Alzheimer disease in the United States (2010-2050) estimated using the 2010 census. Neurology 80, 1778–1783 (2013).

3. Anderson, G. F. & Hussey, P. S. Population Aging : A. (2000).

4. de Meijer, C., Wouterse, B., Polder, J. & Koopmanschap, M. The effect of population aging on health expenditure growth: A critical review. Eur. J. Ageing 10, 353–361 (2013).

5. Deb, A., Thornton, J. D., Sambamoorthi, U. & Innes, K. Direct and indirect cost of managing alzheimer’s disease and related dementias in the United States. Expert Review of Pharmacoeconomics and Outcomes Research (2017) doi:10.1080/14737167.2017.1313118.

6. Langa, K. M. et al. Out-of-Pocket Health Care Expenditures Among Older Americans with Dementia. Alzheimer Dis. Assoc. Disord. 18, 90–98 (2004).

7. Martyr, A. et al. Living well with dementia: A systematic review and correlational meta-analysis of factors associated with quality of life, well-being and life satisfaction in people with dementia. Psychological Medicine (2018) doi:10.1017/S0033291718000405.

8. Medeiros, R. et al. Inflammation: the link between comorbidities, genetics, and Alzheimer’s disease. J. Neuroinflammation (2018) doi:10.1186/s12974-018-1313-3.

9. Doraiswamy, P. M., Leon, J., Cummings, J. L., Marin, D. & Neumann, P. J. Prevalence and impact of medical comorbidity in Alzheimer’s disease. Journals Gerontol. - Ser. A Biol. Sci. Med. Sci. 57, M173–M177 (2002).

10. Xu, W. et al. Meta-analysis of modifiable risk factors for Alzheimer’s disease. J. Neurol. Neurosurg. Psychiatry (2015) doi:10.1136/jnnp-2015-310548.

11. Hersi, M. et al. Risk factors associated with the onset and progression of Alzheimer’s disease: A systematic review of the evidence. Neurotoxicology (2017) doi:10.1016/j.neuro.2017.03.006.

12. Williamson, J., Goldman, J. & Marder, K. S. Genetic aspects of alzheimer disease. Neurologist (2009) doi:10.1097/NRL.0b013e318187e76b.

13. Gatz, M. et al. Heritability for Alzheimer’s disease: The study of dementia in Swedish twins. Journals Gerontol. - Ser. A Biol. Sci. Med. Sci. 52, 117–125 (1997).

14. Cuyvers, E. & Sleegers, K. Genetic variations underlying Alzheimer’s disease: Evidence from genome-wide association studies and beyond. Lancet Neurol. 15, 857–868 (2016).

15. Ridge, P. G., Mukherjee, S., Crane, P. K. & Kauwe, J. S. K. Alzheimer’s disease: Analyzing the missing heritability. PLoS One 8, (2013).

16. Kunkle, B. W. et al. Genetic meta-analysis of diagnosed Alzheimer’s disease identifies new risk loci and implicates Aβ, tau, immunity and lipid processing. Nat. Genet. 51, 414– 430 (2019).

17. Jansen, I. E. et al. Genome-wide meta-analysis identifies new loci and functional pathways influencing Alzheimer’s disease risk. Nat. Genet. 51, 404–413 (2019).

18. Kunkle, B. W. et al. Novel Alzheimer Disease Risk Loci and Pathways in African American Individuals Using the African Genome Resources Panel: A Meta-analysis. JAMA Neurol. 78, 102–113 (2021).

19. Corder, E. H. et al. Gene dose of apolipoprotein E type 4 allele and the risk of Alzheimer’s disease in late onset families. Science (80-.). 261, 921–923 (1993).

20. Rajabli, F. et al. Ancestral origin of ApoE ε4 Alzheimer disease risk in Puerto Rican and African American populations. PLoS Genet. 14, 4–11 (2018).

21. Liu, Y. et al. APOE genotype and neuroimaging markers of Alzheimer’s disease: Systematic review and meta-analysis. J. Neurol. Neurosurg. Psychiatry 86, 127–134 (2015).

22. O’Donoghue, M. C., Murphy, S. E., Zamboni, G., Nobre, A. C. & Mackay, C. E. APOE genotype and cognition in healthy individuals at risk of Alzheimer’s disease: A review. Cortex vol. 104 103–123 (2018).

23. Tzioras, M., Davies, C., Newman, A., Jackson, R. & Spires-Jones, T. Invited Review: APOE at the interface of inflammation, neurodegeneration and pathological protein spread in Alzheimer’s disease. Neuropathol. Appl. Neurobiol. 45, 327–346 (2019).

24. Hostetler, J. Amish Society. (Johns Hopkins University Press, 1993).

25. Jackson, C. E., Symon, W. E., Pruden, E. L., Kaehr, I. M. & Mann, J. D. Consanguinity and blood group distribution in an Amish Isolate. Am. J. Hum. Genet. 20, 522–527 (1968).

26. Agarwala, R., Biesecker, L. G., Tomlin, J. F. & Sch⍰ffer, A. A. Towards a complete North American Anabaptist genealogy: A systematic approach to merging partially overlapping genealogy resources. Am. J. Med. Genet. 86, 156–161 (1999).

27. Lee, W. J., Pollin, T. I., O’Connell, J. R., Agarwala, R. & Schäffer, A. A. PedHunter 2.0 and its usage to characterize the founder structure of the Old Order Amish of Lancaster County. BMC Med. Genet. 11, 68 (2010).

28. Hatzikotoulas, K., Gilly, A. & Zeggini, E. Using population isolates in genetic association studies. Brief. Funct. Genomics 13, 371–377 (2014).

29. D’Aoust, L. N. et al. Examination of candidate exonic variants for association to alzheimer disease in the Amish. PLoS One 10, 1–16 (2015).

30. Pericak-Vance, M. A. et al. Alzheimer’s disease and apolipoprotein e-4 allele in an amish population. Ann. Neurol. 39, 700–704 (1996).

31. Johnson, C. C., Rybicki, B. A., Brown, G. & Jackson, C. E. Prevalence of dementia in the Amish: a three county survey. Am. J. Epidemiol. 138, (1993).

32. Cummings, A. C. et al. Genome-Wide Association and Linkage Study in the Amish Detects a Novel Candidate Late-Onset Alzheimer Disease Gene. Ann. Hum. Genet. 76, 342–351 (2012).

33. Ashley-Koch, A. E. et al. An autosomal genomic screen for dementia in an extended Amish family. Neurosci. Lett. 379, 199–204 (2005).

34. van der Walt, J. M. et al. Maternal lineages and Alzheimer disease risk in the Old Order Amish. Hum. Genet. 118, 115–122 (2005).

35. Waksmunski, A. R. et al. Rare variants and loci for age-related macular degeneration in the Ohio and Indiana Amish. Hum. Genet. 138, 1171–1182 (2019).

36. Hoffman, J. D. et al. Rare complement factor H variant associated with age-related macular degeneration in the Amish. Investig. Ophthalmol. Vis. Sci. 55, 4455–4460 (2014).

37. Sardell, R. J. et al. Heritability of Choroidal Thickness in the Amish. Ophthalmology 123, 2537–2544 (2016).

38. Courtenay, M. D. et al. Mitochondrial Haplogroup X is associated with successful aging in the Amish. doi:10.1007/s00439-011-1060-3.

39. Edwards, D. R. V. et al. Linkage and association of successful aging to the 6q25 region in large Amish kindreds. Age (Omaha). 35, 1467–1477 (2013).

40. Edwards, D. R. V. et al. Successful Aging Shows Linkage to Chromosomes 6, 7, and 14 in the Amish. Ann. Hum. Genet. 75, 516–528 (2011).

41. Chui, T. E. L. Modified Mini Mental State Examination (3MS).

42. Galvin, J. E., Roe, C. M. & Morris, J. C. Evaluation of cognitive impairment in older adults: Combining brief informant and performance measures. Arch. Neurol. 64, 718–724 (2007).

43. Welsh, K. A. et al. The Consortium to Establish a Registry for Alzheimer’s Disease (CERAD). Part V. A normative study of the neuropsychological battery. Neurology 44, 609–609 (1994).

44. Tombaugh, T.N. Trail Making Test A and B: Normative data stratified by age and education. Arch. Clin. Neuropsychol. 19, 203–214 (2004).

45. Multi-Ethnic Genotyping Array Consortium. https://www.illumina.com/science/consortia/human-consortia/multi-ethnic-genotyping-consortium.html.

46. Global Screening Array Consortium. https://www.illumina.com/science/consortia/human-consortia/global-screening-consortium.html.

47. Beecham, G. W. et al. Clinical/Scientific Notes: The Alzheimer’s disease sequencing project: Study design and sample selection. Neurol. Genet. 3, (2017).

48. Kuzma, A. et al. NIAGADS: The NIA Genetics of Alzheimer’s Disease Data Storage Site. Alzheimer’s Dement. (2016) doi:10.1016/j.jalz.2016.08.018.

49. Loh, P.-R. et al. Reference-based phasing using the Haplotype Reference Consortium panel. Nat. Genet. Vol. 48, (2016).

50. Das, S. et al. Next-generation genotype imputation service and methods. Nat. Genet. 48, 1284–1287 (2016).

51. Manichaikul, A. et al. Robust relationship inference in genome-wide association studies. Bioinformatics (2010) doi:10.1093/bioinformatics/btq559.

52. Liang, X. et al. Genomic convergence to identify candidate genes for Alzheimer disease on chromosome 10. Hum. Mutat. 30, 463–471 (2009).

53. Kohli, M. A. et al. Repeat expansions in the C9ORF72 gene contribute to Alzheimer’s disease in Caucasians. Neurobiol. Aging 34, 1519.e5-1519.e12 (2013).

54. Choi, S. W. & O’Reilly, P.F. PRSice-2: Polygenic Risk Score software for biobank-scale data. Gigascience 8, 1–6 (2019).

55. R Development Core Team, R. R: A Language and Environment for Statistical Computing. R Foundation for Statistical Computing (2011). doi:10.1007/978-3-540-74686-7.

56. Johnson, C. C. et al. Cognitive impairment in the Amish: A four county survey. Int. J. Epidemiol. (1997) doi:10.1093/ije/26.2.387.

