## Supplemental Information for "The genetic architecture of Alzheimer disease risk in the Ohio and Indiana Amish"

**Supplemental Table 1. Kinship coefficient of Amish subpopulations by primary recruitment site and cognitive impairment status.**

| Group | Mean Kinship Coefficient |
|---|---|
| All Amish adults | 0.003703 |
| Primary site: Ohio | 0.006157 |
| Primary site: Indiana | 0.005290 |
| Cognitively Normal | 0.003909 |
| Cognitively Impaired | 0.003793 |

**Supplemental Figure 1. Age distribution of Amish and non-Amish populations.** We observe that the age distributions of the populations are not identical and include many younger individuals, especially in the non-Amish comparison group. This justifies the use of an analysis after age subset in the case of an age-related disease, such as Alzheimer disease.

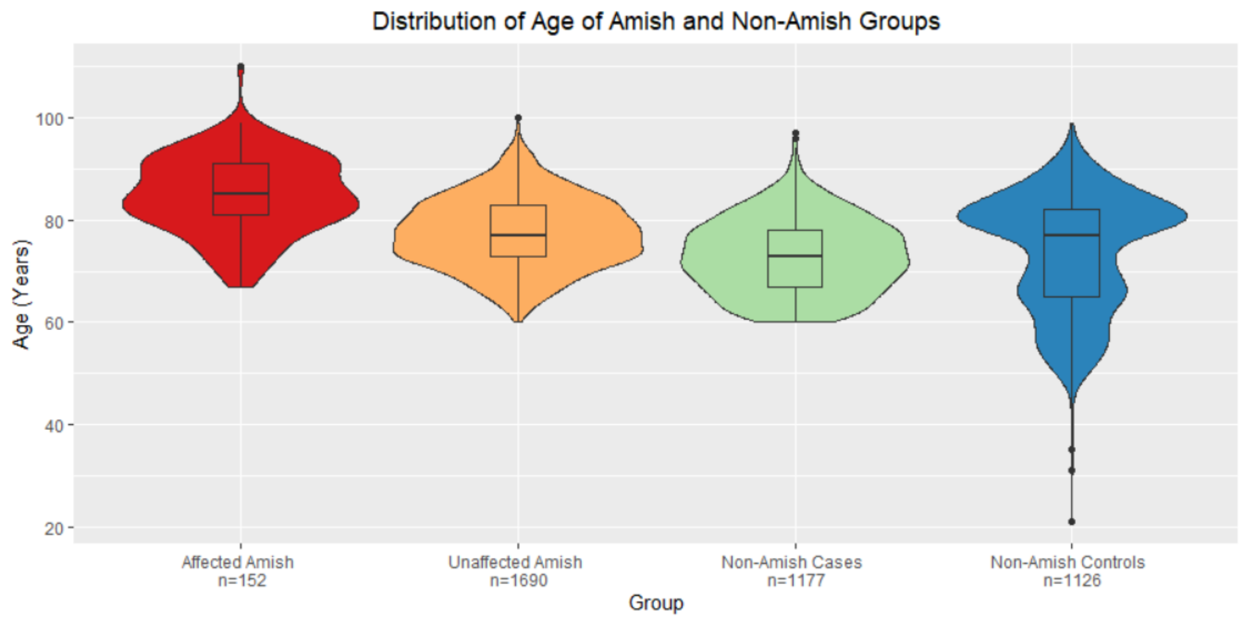


**Supplemental Figure 2. PRS distributions in Amish with cognitive impairment as primary outcome.** We observe that the Jansen et al. (2019)[17] polygenic risk score is capable of distinguishing between Amish affected by cognitive impairment and unaffected by cognitive impairment (p=0.002). *Abbreviations: CI = cognitively impaired*.

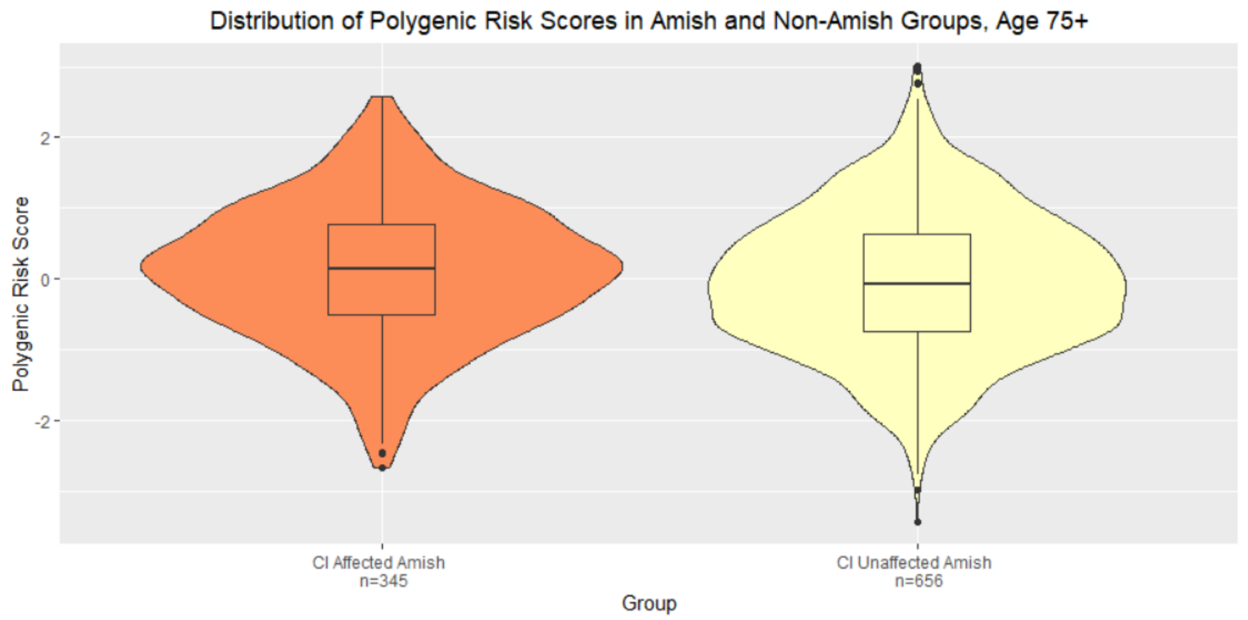
Distribution of Polygenic Risk Scores in Amish and Non-Amish Groups, Age 75+
Polygenic Risk Score
2
0
-2
CI Affected Amish
n=345
CI Unaffected Amish
n=656
Group
